# Genomic Insights into the Shared and Distinct Genetic Architecture of Cognitive Function and Schizophrenia

**DOI:** 10.1101/2023.11.13.23298348

**Authors:** Olivia Wootton, Alexey A. Shadrin, Thomas Bjella, Olav B. Smeland, Dennis van der Meer, Oleksandr Frei, Kevin S O’Connell, Torill Ueland, Ole A. Andreassen, Dan J. Stein, Shareefa Dalvie

**Author notes:** Corresponding author: Olivia Wootton, UCT Department of Psychiatry and Mental Health, Groote Schuur Hospital, Anzio Road, Observatory, 7925, Cape Town, South Africa.

## Abstract

Cognitive impairment is a major determinant of functional outcomes in schizophrenia, and efforts to understand the biological basis of cognitive dysfunction in the disorder are ongoing. Previous studies have suggested genetic overlap between global cognitive ability and schizophrenia, but further work is needed to delineate the shared genetic architecture. Here, we apply genomic structural equation modelling to identify latent cognitive factors capturing genetic liabilities to 12 cognitive traits measured in the UK Biobank (UKB). We explore the overlap between latent cognitive factors, schizophrenia, and schizophrenia symptom dimensions using a complementary set of statistical approaches, applied to data from the latest schizophrenia genome-wide association study (Ncase = 53,386, Ncontrol = 77,258) and the Thematically Organised Psychosis study (Ncase = 306, Ncontrol = 1060). We identified three broad factors (visuo-spatial, verbal analytic and decision/reaction time) that underly the genetic correlations between the UKB cognitive tests. Global genetic correlations showed a significant but moderate negative genetic correlation between each cognitive factor and schizophrenia. Local genetic correlations implicated unique genomic regions underlying the overlap between schizophrenia and each cognitive factor. We found evidence of substantial polygenic overlap between each cognitive factor and schizophrenia but show that most loci shared between the latent cognitive factors and schizophrenia have unique patterns of association with the cognitive factors. Biological annotation of the shared loci implicated gene-sets related to neurodevelopment and neuronal function. Lastly, we find that the common genetic determinants of the latent cognitive factors are not predictive of schizophrenia symptom dimensions. Overall, these findings inform our understanding of cognitive function in schizophrenia by demonstrating important differences in the shared genetic architecture of schizophrenia and cognitive abilities.

## Introduction

A global deficit in cognitive function is characteristic of schizophrenia^1,2^ with evidence indicating greater impairment for specific cognitive domains such as executive function, attention, episodic memory, and motor speed, compared to others.^3,4^ Cognitive impairment is a major determinant of functional outcomes in schizophrenia^5^ yet most existing therapies for schizophrenia do not address cognitive symptoms.^6^ Thus, there has been increasing interest in identifying the biological basis of cognitive dysfunction in schizophrenia in order to facilitate the identification of novel therapeutic targets.^6^

Both schizophrenia and cognitive ability are heritable, with estimates from twin and family studies of 0.8 for schizophrenia^7^ and 0.5 for cognitive ability^8^. Previous research has demonstrated impaired cognitive performance in first degree relatives of people with schizophrenia,^9,10^ providing further evidence that there is a genetic contribution to cognitive dysfunction in the disorder. Molecular genetic studies have identified rare and common genetic variants that are associated with both schizophrenia and cognitive function, implicating genes involved in neurodevelopment, synaptic integrity, and neurotransmisison.^11,12^ However, findings from studies investigating the association between genetic liability for schizophrenia and cognitive impairment in individuals with schizophrenia have been inconsistent. Some studies have found that a polygenic risk score (PRS) for schizophrenia is negatively associated with cognitive ability^13,14^ while others have found no association^15–17^.

One possible explanation for the inconsistent results is that most previous studies have focused on the association between the genetic determinants of schizophrenia and a single measure of general cognitive ability, such as Spearman’s *g* factor. Given that performance across cognitive domains is known to be differentially affected by schizophrenia, the use of a broad measure of cognitive ability may prevent a more detailed examination of the genetic contributions to cognitive impairment in the disorder. Additionally, schizophrenia is a clinically heterogenous disorder^18^ and there is evidence to suggest that unique biological processes contribute to the various schizophrenia symptom dimensions^19,20^. While it is plausible that the degree of genetic overlap with cognitive function will differ across the schizophrenia symptom dimensions, few genetic studies have taken a dimensional approach to assessing the relationship.

Here, we take a nuanced approach to investigating the shared genetic determinants between cognitive function and schizophrenia. We apply Genomic Structural Equation Modelling (SEM) to derive latent factors corresponding to broad dimensions of cognitive function from 12 cognitive traits measured in the UK Biobank (UKB). We use novel statistical approaches to identify genetic variants shared between the dimensions of cognitive function and schizophrenia. Lastly, we explore whether the phenotypic distinction between cognitive dysfunction and other schizophrenia symptoms may be explained by differences in the underlying genetic architecture.

## Materials and Methods

### Sample description

This study used data from the UKB, a large-scale biomedical database with genotype and phenotype data for approximately 500,000 people.^21^ Data for this study was obtained under accession number 27412. Our analysis uses genotype and cognitive data for individuals with a self-reported ethnicity of “white British” or “white non-British”. Cognitive tests included in this study were completed during the baseline assessment, and later follow-up assessments. Sample sizes for each cognitive measure vary (*n* = 28,156 – 436,853; Figure 1) and participants’ ages range from 40-70 years at baseline and 45-75 years at later assessments.

**Figure 1.**
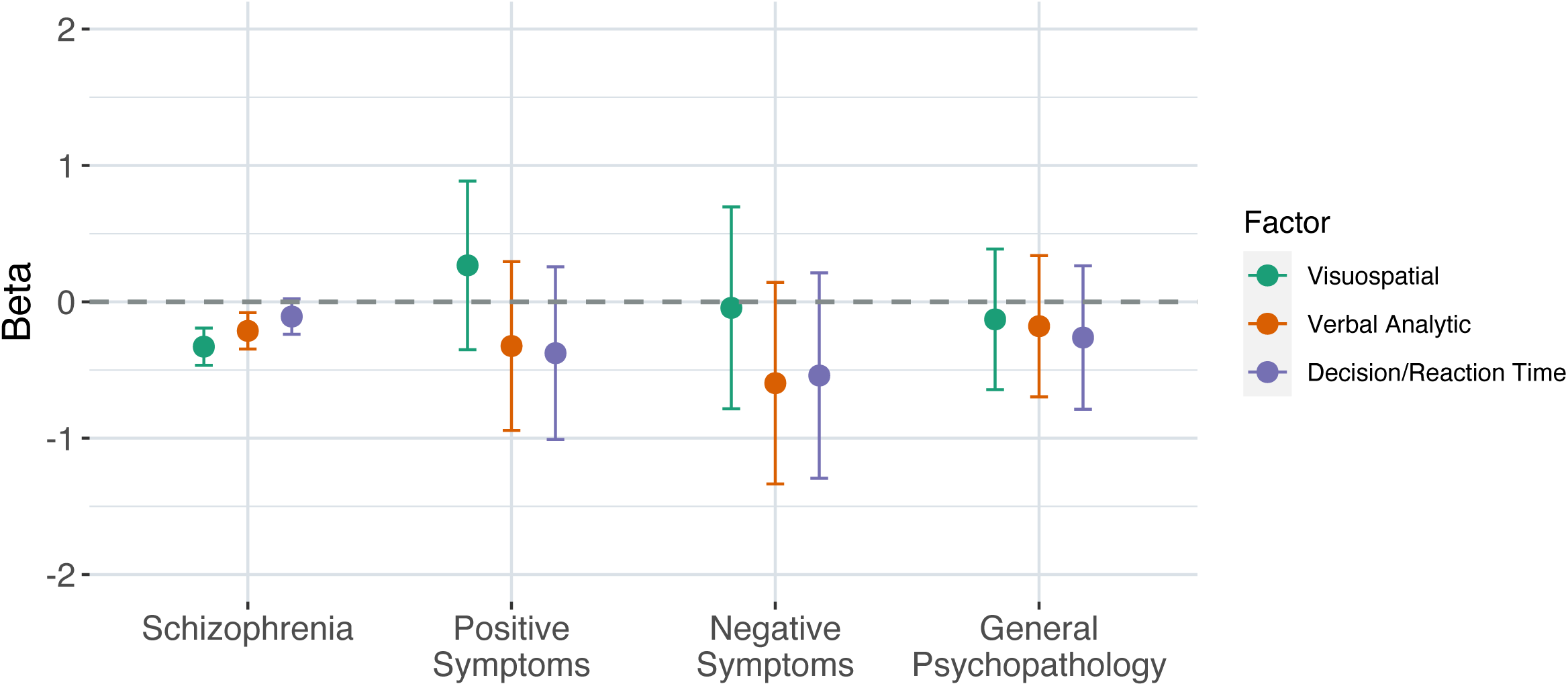
Overview of univariate GWAS for UKB cognitive traits. Bar chart illustrating the sample size, number of genome-wide significant loci, and SNP-based heritability estimates for univariate GWAS of 12 cognitive traits from the UK Biobank. Traits are colour-coded according to the three-factor model derived using Genomic Structural Equation Modelling. Pairs Match, Pairs Matching; TMT A, Trail Making Test-Part A; TMT B, Trail Making Test-Part B; SDS, Symbol Digit Substitution; Tower, Tower Rearranging; Matrix, Matrix Pattern Completion; PAL, Paired Associate Learning; Fluid Int, Fluid Intelligence; Num Mem, Numeric Memory; Pro Mem, Prospective Memory; Mean RT, Mean Reaction Time; RTV, Reaction Time Variability.

### Definition of the cognitive phenotypes

This study included 12 cognitive measures that were derived from cognitive tests administered as part of the baseline and follow up assessments for the UKB. The four cognitive tests that were administered at baseline are Fluid Intelligence, Reaction Time, Numeric Memory, and Pairs Matching Test. The six tests that were administered during a follow up assessment are Prospective Memory, Matrix Pattern Completion, Paired Associate Learning (PAL), UKB Symbol Digit Substitution, Tower Rearranging, UKB Trail Making Test – Part A and B (TMT-A and TMT-B). All cognitive tests were fully automated and were designed to be administered with minimal supervision. A detailed description of each cognitive test is provided in the Supplementary Note.

### Genome-wide association analyses

Version 3 of the UKB genetic data was used for this study. Genotyping, imputation, and central quality control procedures for the UKB genotypes are described in detail elsewhere.^22^ Univariate genome-wide association studies (GWAS) for each cognitive phenotype was conducted using the REGENIE tool^23^, which consists of two steps. For step 1, polygenic predictors are calculated by fitting a whole genome regression model to genotype data. Prior to conducting step 1, the following quality control filters were applied to the UKB genotype calls: removal of individuals with > 10% missing genotype data, removal of SNPs with > 10% genotype missingness, removal of SNPs failing the Hardy-Weinberg equilibrium tests at *p* = 1 x 10^-15^, and removal of SNPs with a minor allele frequency (MAF) < 1%. After quality control, 581,299 variants remained for inclusion in step 1 of the analysis. For step 2, a linear regression model is used to test for phenotype-genotype associations using imputed genotype data, conditional upon the predictions of the model from step 1. Variants with an INFO score < 0.8 and MAC < 20 were excluded from this step, leaving a maximum of 20,241,796 variants for analysis. Sex, age, age^2^, age by sex interaction, assessment centre, genotype array, and the first 40 genetic principal components were included as covariates in each GWAS.

### Estimations of SNP-based heritability and genetic correlations

The SNP-based heritability (*h^2^*_SNP_) for each cognitive phenotype from the UKB was estimated using linkage disequilibrium score regression (LDSC)^24,25^. LDSC was also used to estimate the genetic correlations between the 12 cognitive phenotypes.

### Genomic structural equation modelling

We conducted exploratory and confirmatory factor analysis of the 12 UKB cognitive phenotypes. First, the multivariable extension of LDSC employed in Genomic SEM was used to derive a genetic covariance matrix (S) and sampling covariance matrix (V). Next, exploratory factor analysis (EFA) with promax rotation was conducted on the standardized S matrix using the R package, *stats*^26^. Results from the EFA were used to guide confirmatory factor analysis (CFA) for a one, two-, and three-factor model. CFA was performed using Genomic SEM and standardized factor loadings of > 0.4 were retained for CFA. Model fit for each factor model was assessed using recommended fit indices: standardized root mean square residual (SRMR), model χ2 statistic, Akaike Information Criterion (AIC), and Comparative Fit Index (CFI). Model fit was considered acceptable for CFI values ≥ 0.90 and SRMR values ≤ 10.^27^ A three-factor solution demonstrated superior model fit to a one- or two-factor solution and was selected for subsequent analysis.

### Multivariate GWAS in Genomic SEM

Following identification of the confirmatory factor model that best explained the genetic covariance structure among the UKB cognitive phenotypes, Genomic SEM was used to estimate the individual SNP associations with each latent factor in the model. As the cross-trait intercepts estimated by multivariable LDSC account for sample overlap, SNP association estimates derived using Genomic SEM are robust to varying and unknown degrees of sample overlap across the contributing univariate GWAS.^27–29^ The multivariate GWAS was conducted using summary statistics for the univariate GWAS for each cognitive phenotype. Prior to conducting the multivariate GWAS, effect alleles were aligned across univariate GWAS and beta coefficients were standardized. Summary statistics for input into the multivariate GWAS were restricted to SNPs that were present for all 12 cognitive phenotypes and present in the 1000 Genomes Project Phase 3 release European reference panel^30^. After filtering, 8,041,728 SNPs remained for inclusion in the multivariate GWAS. The effective sample size for each latent factor was calculated using methodology described in a previous publication^31^.

### Estimation of genetic overlap between cognitive factors and schizophrenia

We explored the genetic overlap between the three latent cognitive factors and schizophrenia using summary statistics from the multivariate GWASs and for participants of European ancestry in the latest PGC Schizophrenia GWAS (Ncase = 53,386, Ncontrol = 77,258)^32^.

First, global genetic correlations between the latent cognitive factors and schizophrenia were estimated using LDSC^24,25^. Bivariate MiXeR was used to estimate the number of phenotype-specific and shared causal variants between each cognitive factor and schizophrenia.^33^ A bivariate Gaussian mixture model with four components was constructed using summary statistics for each cognitive factor and schizophrenia. The four components of the model represent 1) SNPs with a null effect for both phenotypes, 2 and 3) SNPs with a non-null effect for either the first or second phenotype, and 4) SNPs with a non-null effect for both phenotypes. Model fit was evaluated by the AIC.

Next, local genetic correlations between the latent cognitive factors and schizophrenia were estimated using Local Analysis of [co]Variant Association (LAVA)^34^. For each phenotype, LAVA was used to estimate the genetic variance across 2,495 semi-independent genetic loci of approximately equal size (∼1 Mb) defined by Werme et al.^34^ Loci with a significant local SNP based heritability (α = 0.05/2,495 loci; *p* < 2 x 10^-5^) for each phenotype were included in the bivariate analysis. LAVA estimates local genetic correlations for each phenotype pair by constructing a matrix of local genetic covariance for each locus using the method of moments.

Lastly, we conducted conjFDR analysis^35,36^ to identify individual SNPs that were jointly associated with each latent cognitive factor-schizophrenia pair. The conjFDR method is an extension of the condFDR approach^35,37^. For each latent cognitive factor-schizophrenia pair, we used SNP associations with the latent cognitive factor to re-rank the test statistics and recalculate the significance of the SNP associations with schizophrenia. We then reversed the phenotypes and re-calculated the strength of a SNP association with each cognitive factor conditional on the SNP association with schizophrenia. Next, conjFDR analysis was used to estimate the likelihood, represented as a conjFDR value, that a SNP has a non-null association with both phenotypes in a phenotype pair. A conjFDR value < 0.05 was considered significant.

### Functional Annotation

We used standard Functional Mapping and Annotation of Genome-wide Association Studies (FUMA) definitions to define genomic loci, lead SNPs, independent significant SNPs and candidate SNPs by clumping the conjFDR output for each latent cognitive factor-schizophrenia pair at an FDR of < 0.05. Next, we used Bedtools v2.27.1^38^ with default parameters to identify significant loci that were unique to a latent cognitive factor-schizophrenia pair. We mapped genes to the unique significant loci using data provided by Ensembl^39^ based on the GRCh38 reference genome. We used the GENE2FUNC function in FUMA to test for enrichment of the identified genes for each latent cognitive factor-schizophrenia pair in gene sets obtained from MsigDB v7.0^40^. The Benjamini-Hochberg correction for multiple testing was applied per category of gene-sets.

### Polygenic prediction of schizophrenia symptom dimensions

For polygenic score (PGS) analyses, the target dataset comprised 306 individuals with schizophrenia and 1060 controls of European ancestry from the Norwegian *Thematically Organised Psychosis* (TOP) Study^41^. For individuals with schizophrenia, symptoms were measured using the positive, negative, and general psychopathology subscales of the Positive and Negative Syndrome Scale (PANSS)^42^. Mean scores for the PANSS in the TOP sample were 14.37 (SD = 5.29) for the positive scale, 15.30 (SD = 6.26) for the negative scale, and 12.95 (SD = 4.33) for the general psychopathology scale. The recruitment and genotyping procedures for the TOP study are described in the Supplementary Note. A PGS for each of the latent cognitive factors was calculated from the effect size estimates from the multivariate GWAS summary statistics using PRS-CS-auto^43^. PRS-CS-auto is a Bayesian polygenic prediction method that estimates posterior effect sizes of SNPs by placing a continuous shrinkage prior on SNP effect sizes and incorporating information from an external LD reference panel. PRS-CS-auto automatically estimates the global shrinkage prior from the discovery dataset and does not require a validation dataset.^43^ In the present study, the 1000 Genomes Phase 3 release European sample^30^ was used as the LD reference panel. SNPs with a minor allele frequency < 0.01 were excluded from the analysis, which left 964,446 SNPs for calculation of the PGS. Linear regression models were used to test the association between PGS for each latent cognitive factor with schizophrenia and the three schizophrenia symptom dimensions. Age, sex, and the first 20 genetic principal components were included as covariates in the model. Phenotypic variance explained by the PGS (Nagelkerke’s pseudo-R^2^ for schizophrenia diagnosis and R^2^ for PANSS subscale scores) was estimated as the difference between the R^2^ of the full regression model (PGS and covariates) and the R^2^ of the null model (covariates only). The Bonferroni correction was applied to account for multiple testing (α = 0.05/12 polygenic scores; *p* < 4.17 x 10^-3^).

## Results

### Genome-wide association analyses of UKB cognitive traits

We conducted univariate GWASs of 12 cognitive traits from the UKB. The number of genome-wide significant loci identified for each cognitive trait ranged from 0 to 87 with the highest number of genome-wide significant loci observed for mean reaction time and no significant loci for PAL and TMT-B (Figure 1; Supplementary Table 1). The LD score regression intercept for all univariate GWAS was approximately 1 (range: 0.99 - 1.02), consistent with minimal inflation of the test statistic due to population stratification (Supplementary Table 1).

### Estimations of SNP-based heritability and genetic correlations

The *h^2^*_SNP_ for each cognitive trait and the genetic correlations between cognitive traits were estimated using LDSC. Estimates for *h^2^*_SNP_ ranged from 3 - 22% (Figure 1; Supplementary Table 1). We observed positive genetic correlations between all cognitive traits (range 0.08 – 0.89, mean = 0.49, SD = 0.2; Figure 2A). All genetic correlations were significant (α = 0.05/66 pairs of traits; *p* < 7.58 x 10^-4^) with the exception of the correlations between mean reaction time and PAL (r_g_ = 0.11, SE = 0.04, *p* = 5.3 x 10^-3^), mean reaction time and numeric memory (r_g_ = 0.07, SE = 0.03, *p* = 1.74 x 10^-2^), and TMT-A and PAL (r_g_ = 0.25, SE = 0.09, *p* = 6.2 x 10^-3^). We applied a hierarchal clustering algorithm to the genetic correlation matrix and identified three distinct clusters of cognitive phenotypes. The first cluster included Pairs Matching, TMT-A, TMT-B, Symbol Digit Substitution and Tower Rearranging, the second cluster included Fluid Intelligence, Numeric Memory, Prospective Memory, Matrix Reasoning, and PAL, and the third cluster comprised of mean reaction time and reaction time variability (Figure 2B).

**Figure 2.**
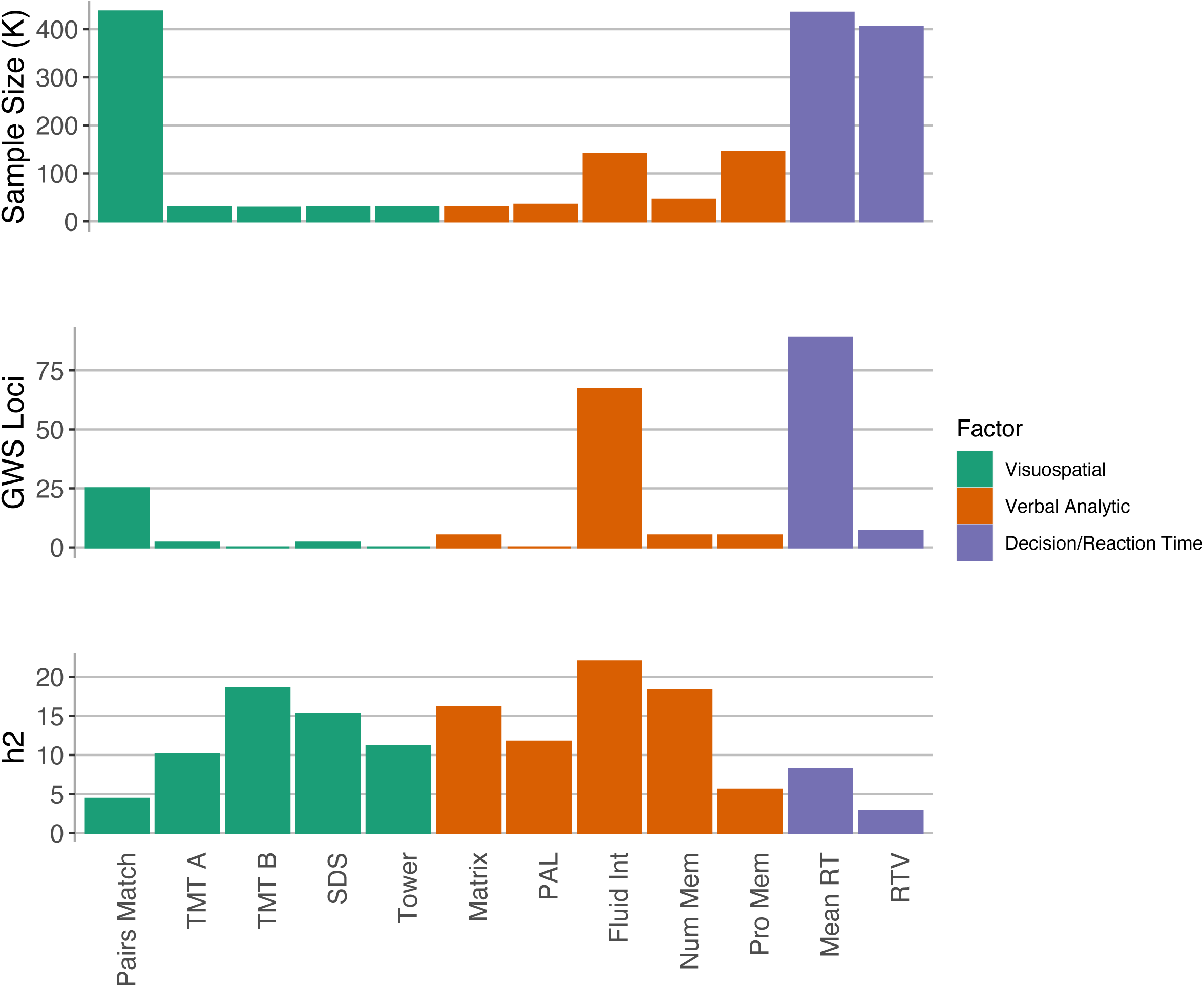
Multivariate structure of 12 cognitive traits from the UK Biobank. (**A**) Heatmap showing genetic correlations estimated using LDSC. (**B**) Path diagram showing standardized results from the correlated three-factor model. Circles represent latent variables that are inferred from the data. Single-headed arrows depict regression relationships with the arrows pointing from the independent variables to the dependent variables. Two-headed arrows represent covariance relationships between variables or the residual variance of a variable if the arrow connects the variable to itself. Standard errors of the estimates are in parenthesis. Pairs Match, Pairs Matching; TMT A, Trail Making Test-Part A; TMT B, Trail Making Test-Part B; SDS, Symbol Digit Substitution; Tower, Tower Rearranging; Matrix, Matrix Pattern Completion; PAL, Paired Associate Learning; Fluid Int, Fluid Intelligence; Num Mem, Numeric Memory; Pro Mem, Prospective Memory; Mean RT, Mean Reaction Time; RTV, Reaction Time Variability.

### Genomic structural equation modelling

We modeled the genetic covariance matrix for the 12 UKB cognitive traits using Genomic SEM. First, we fit a single common factor model in which the single latent factor represented a genetic *g* factor. Model fit was suboptimal for the single common factor model (chi-square, χ2(54) = 2106.75, AIC = 2154.75, CFI = 0.71, SRMR = 0.12; Supplementary Table 2; Supplementary Figure 1). Next, we explored whether a correlated two- or three-factor model closely approximated the observed genetic covariance matrix. Consistent with the results from the hierarchal clustering of the genetic correlation matrix, we found that a three correlated factors model fit the data best (chi-square, χ2(49) = 320.72, AIC = 378.72, CFI = 0.96, SRMR = 0.07; Figure 2B; Supplementary Table 2). In the three-factor model, factor 1 and factor 2 exhibit the highest genetic correlation among the cognitive factors (r_g_ = 0.65, SE = 0.02, *p* = 2.59 x 10^-215^) and these factors may be conceptualized as capturing cognitive traits related to the broad cognitive ability, fluid reasoning^44^. Factor 1 is primarily defined by cognitive phenotypes that relate to visuospatial aspects of fluid reasoning and includes Pairs Matching, TMT-A, TMT-B, Symbol Digit Substitution and Tower Rearranging. Factor 2 largely captures measures that assess the verbal analytic component of fluid reasoning and is defined by Fluid Intelligence, Numeric Memory, Prospective Memory, Matrix Reasoning, and PAL. Factor 3 is characterized by cognitive phenotypes related to the broad cognitive ability, “decision/reaction time/speed”^44^. The cognitive measures, mean reaction time and reaction time variability load on the third factor, which is less correlated with factor 1 (r_g_ = 0.37, SE = 0.03, *p* = 2.75 x 10^-42^) and factor 2 (r_g_ = 0.29, SE = 0.02, *p* = 3.44 x 10^-37^).

### Multivariate GWAS

We conducted multivariate GWASs of the three latent cognitive factors using Genomic SEM. The effective sample size ranged from 160,729 for factor 2 (verbal analytic reasoning) to 637,271 for factor 1 (visuospatial processing) (Supplementary Table 3). Substantial inflation of the test statistic was observed for all latent cognitive factors (Supplementary Figure 2) however LD score regression intercepts were 1, suggesting that test-statistic inflation reflects high polygenicity and not other sources of bias (Supplementary Table 3).

### Estimation of genetic overlap between cognitive factors and schizophrenia

We found a significant negative genetic correlation between all three latent cognitive factors and schizophrenia using LDSC (Supplementary Table 4). Bivariate MiXeR demonstrated substantial polygenic overlap between each latent cognitive factor and schizophrenia, beyond that captured by estimates of genetic correlation (Supplementary Figure 3). Of the 9,600 variants predicted to influence schizophrenia, almost all variants were also predicted to influence the latent cognitive factors. Notably, factor 1 (visuospatial processing) demonstrated the greatest negative global genetic correlation with schizophrenia (rg = -0.38, SE = 0.025, p = 9x10^-52^) and the greatest percentage of shared variants (63%) with a discordant effect between a latent cognitive factor and schizophrenia (Supplementary Figure 3).

We employed LAVA to explore regional patterns of genetic correlation between the latent cognitive factors and schizophrenia. The number of genetic regions that were significantly heritable (*p* < 2.00x10^-5^) for schizophrenia and a latent cognitive factor ranged from 149 - 170 (Supplementary Table 5). Among the significantly heritable regions, we found the number of regions with a significant genetic correlation (α = 0.05/number of significantly heritable regions for both traits) between schizophrenia and a latent cognitive factor ranged from 6-21. Most significant local genetic correlations were negative (Supplementary Figure 4; Supplementary Table 6) with the exception of a positive correlation between schizophrenia and factor 2 (verbal analytic reasoning) at two loci [chr 3: 47588462-50387742, chr 12: 77800464-79315178] (Supplementary Table 6).

As shown in Figure 3, conjFDR analysis demonstrated that schizophrenia shares loci with cognitive factor 1 (N=93), cognitive factor 2 (N=267), and cognitive factor 3 (N=175). Consistent with lowest degree of polygenic overlap demonstrated by bivariate MiXeR analysis, factor 1 shared the lowest number of loci with schizophrenia. Among the shared associations with schizophrenia, 46 were unique to cognitive factor 1, 189 were unique to factor 2, and 113 were unique to factor 3.

**Figure 3.**
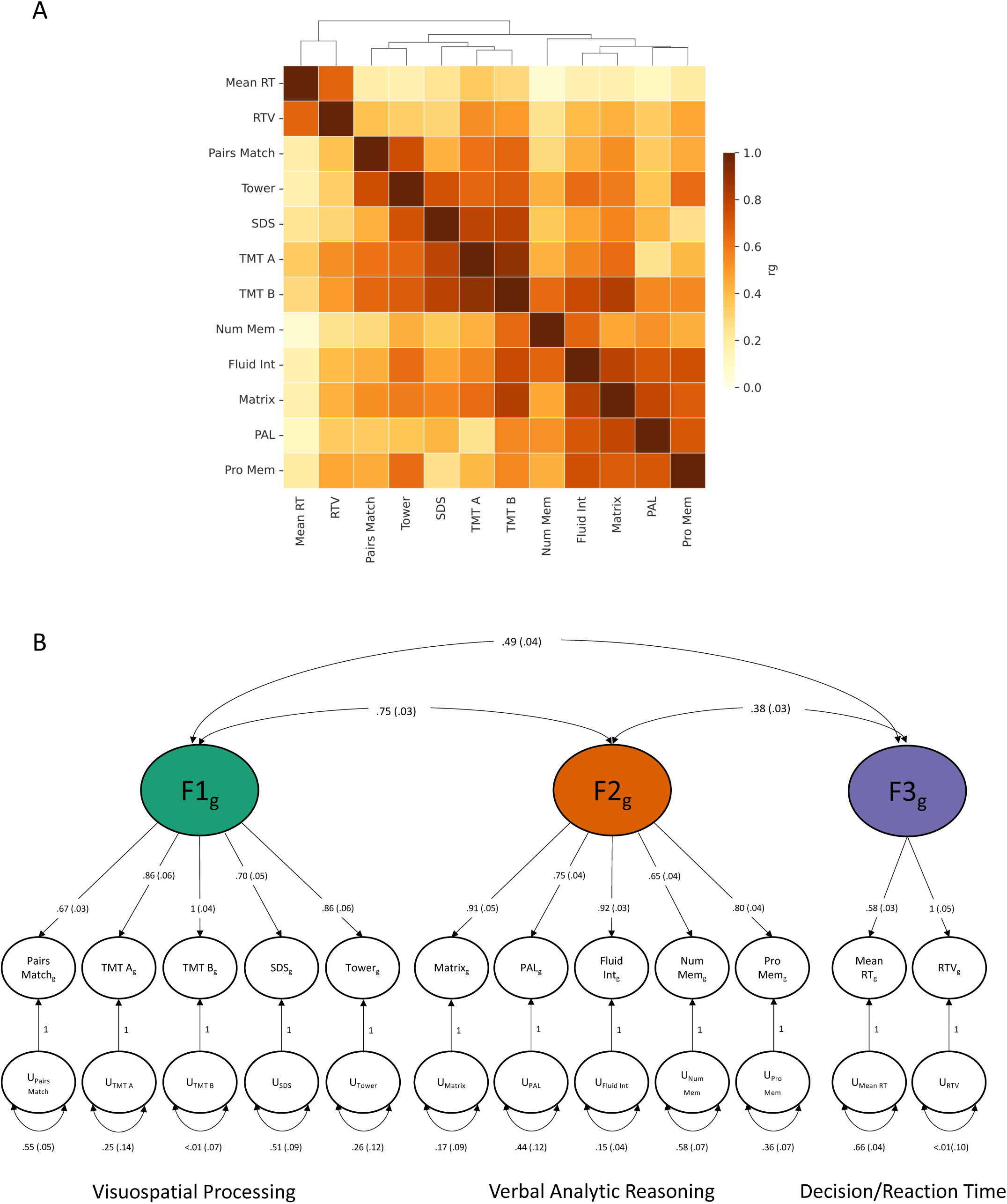
Discovery of loci jointly associated with each latent cognitive factor and schizophrenia. Manhattan plots showing the -log_10_ transformed conjFDR values for a joint association with a latent cognitive factor and schizophrenia (SCZ) for each SNP (y-axis) plotted against chromosomal position (x-axis). The dotted line indicates the significance threshold (conjFDR < 0.05). Lead SNPs are outlined in black and independent significant SNPs are represented by larger circles.

### Functional annotation of identified loci

The genes mapped to unique significant loci for each latent cognitive factor-schizophrenia pair are listed in Supplementary Tables 7-9. For the unique significant loci shared between cognitive factor 1 (visuospatial processing) and schizophrenia, gene-set analysis for cellular components demonstrated significant results for the synapse (FDR = 1.39 x 10^-2^), the synaptic membrane (FDR = 3.05 x 10^-2^), the postsynaptic membrane (FDR = 3.05 x 10^-2^), neurons (FDR = 3.05 x 10^-2^), and neuron projections (FDR = 4.80 x 10^-2^) and no significant gene-sets for biological processes (Supplementary Table 10). For unique significant loci associated with cognitive factor 2 (verbal analytic reasoning) and schizophrenia, gene-set analysis revealed significant results for genes involved in axon guidance (FDR = 2.02 x 10^-2^) and cell adhesion via plasma adhesion molecules (FDR = 1.49 x 10^-3^) (Supplementary Table 11). Lastly, the genes mapped to unique significant loci associated with latent cognitive factor 3 (decision/reaction time) and schizophrenia were significantly enriched for two gene-sets involved in neuronal development: regulation of neuron differentiation (FDR = 4.22 x 10^-2^) and regulation of neuron projection development (FDR = 4.22 x 10^-2^) (Supplementary Table 12).

### Polygenic prediction of schizophrenia symptom dimensions

We created PGS for the three latent cognitive factors and tested the ability of each cognitive factor-PGS to predict schizophrenia and schizophrenia symptom domains in individuals from the TOP study. There was a significant association between schizophrenia diagnosis and the PGS for latent cognitive factor 1, visuospatial processing, (*R*^2^ = 0.026, *p* = 2.48 x 10^-6^) and latent cognitive factor 2, verbal analytic reasoning (*R*^2^ = 0.011, *p* = 1.80 x 10^-3^) (Figure 4). There were no significant associations found between any of the PGS for the latent cognitive factors and schizophrenia symptom dimensions (Figure 4; Supplementary Table 13).

**Figure 4.**
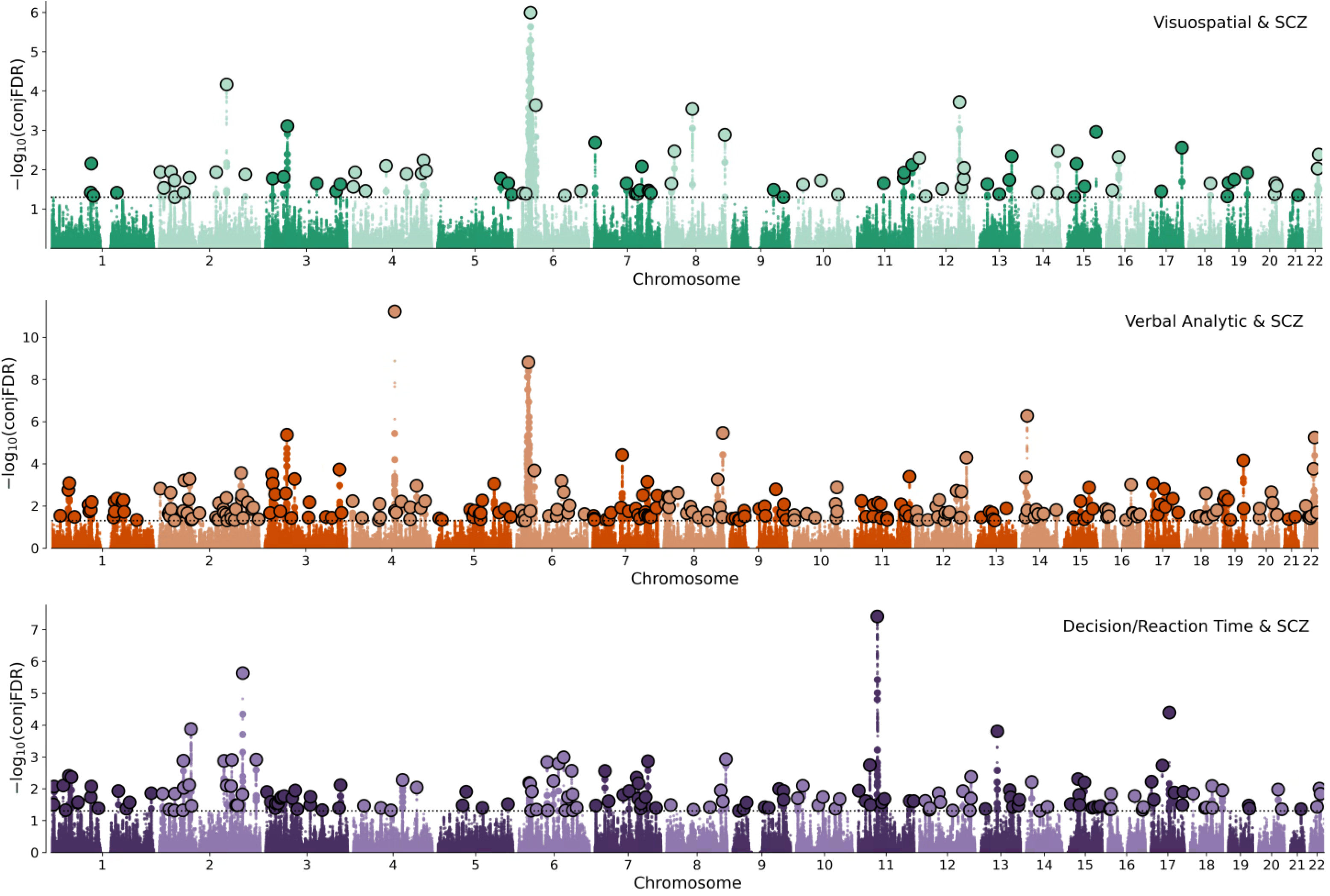
Association of polygenic scores for each latent cognitive factor with schizophrenia and three schizophrenia symptom dimensions. Associations of polygenic scores (PGS) for each latent cognitive factor with schizophrenia, and schizophrenia symptom dimensions (negative, positive, and general psychopathology) in individuals from the TOP study. Point estimates for beta values are shown with 95% confidence intervals. The dotted line represents a null model.

## Discussion

In this study, we explored the common genetic determinants shared between schizophrenia and cognitive function using three latent factors that captured the genetic covariance structure of 12 cognitive measures from the UKB. We found evidence of substantial polygenic overlap between schizophrenia and the genetically determined latent cognitive factors. All three latent cognitive factors exhibited a negative genetic correlation with schizophrenia that was largely consistent for both global and local patterns of genetic correlation. We identified loci jointly associated with schizophrenia and the latent cognitive factors and biological annotation of the shared loci implicated genes involved in the development and functioning of the central nervous system. Additionally, we demonstrated that PGS for the latent cognitive factors were not predictive of schizophrenia symptoms, suggesting distinctions in the underlying genetic architecture of cognitive function and phenotypic dimensions in schizophrenia.

We applied Genomic SEM to GWAS summary statistics for 12 cognitive traits in the UKB and found that a three-factor model best explained the genetic correlations between the cognitive traits. This is consistent with findings from a previous study that applied structural equation modeling to phenotypic data from the UKB cognitive assessments and found that a three-factor solution fit the data best.^45^ The three cognitive factors identified in the current study may be characterized using the framework provided by the Cattell-Horn-Carrol (CHC) theory of human cognitive abilities, which proposes a three-stratum model of human intelligence.^44^ ^46^ Consistent with the relatively high genetic correlation between factor 1 and factor 2, the first two latent cognitive factors appear to capture the same broad cognitive ability, fluid reasoning (*Gf*). However, factor 1 is largely defined by cognitive traits that capture visuospatial processing whereas factor 2 is defined by cognitive tests that measure verbal analytic reasoning. Factor 3 is less genetically correlated with the first two factors and measures a distinct cognitive ability, decision/reaction time/speed (*Gt).* The convergence of the genetic and phenotypic factor structures for the UKB cognitive traits suggests that the existing phenotypic structure is rooted in the genetic underpinnings of the traits.

Consistent with the literature, all three latent cognitive factors demonstrated a significant negative genetic correlation with the diagnosis of schizophrenia with the greatest negative correlation observed between the visuospatial factor (factor 1) and schizophrenia. The genetic correlation between the visuospatial factor and schizophrenia is of greater magnitude than that reported for general cognitive ability and schizophrenia (r_g_ = -0.23)^47^. This finding suggests that we may be missing unique patterns of genetic association between schizophrenia and cognitive abilities when using a composite measure of cognitive function. Patterns of local genetic correlation between the latent cognitive factors and schizophrenia were generally consistent with global genetic correlations. These findings indicate that, in general, genetic liability for schizophrenia has a negative effect on cognitive abilities and that the phenotypic relationship between schizophrenia and cognitive impairment has a genetic basis. LAVA analysis revealed that most significantly heritable regions demonstrated a negative association between the cognitive factors and schizophrenia and that most significant regional genetic correlations were negative. One of the regions that showed a positive genetic correlation between the verbal analytic factor and schizophrenia was located on chromosome 3 (Chr3p21). This region is enriched for genes that have been associated with intelligence^48^ and general cognitive ability^47^ in previous GWAS. Further research is required to explore the relationship between genes in this region and cognitive function in schizophrenia.

Results from bivariate MiXeR and conjFDR analysis converged with each cognitive factor demonstrating substantial polygenic overlap with schizophrenia. Notably, the visuospatial factor demonstrated the greatest genome-wide genetic correlation with schizophrenia despite conjFDR showing that this factor shared the lowest number of loci with schizophrenia. These findings demonstrate that a large genetic correlation does not necessarily correspond to the greatest overlap in genetic architecture. Instead, these findings imply that the variants that are shared between the visuospatial factor and schizophrenia demonstrate a more consistent direction of effect for the two traits than those shared between the other latent cognitive factors and schizophrenia. The conjFDR analysis showed that the majority of loci found to be jointly associated with each latent cognitive factor-schizophrenia pair were unique to the pair. Given the almost complete overlap between the genetic determinants of each cognitive factor and schizophrenia demonstrated by bivariate MiXeR, the difference in significant loci shared between each cognitive factor and schizophrenia is unlikely to reflect a unique set of variants associated with each cognitive factor. Instead, this result indicates that despite all cognitive factors sharing most causal variants, the magnitude and potentially direction of effects of these shared variants vary between the latent cognitive factors. We annotated the unique significant loci for each latent cognitive factor-schizophrenia pair and conducted gene-set enrichment analysis to explore putative biological mechanisms underlying the association between cognitive abilities and schizophrenia. Gene-set enrichment analysis implicated distinct gene-sets for each latent cognitive factor-schizophrenia pair but converged on processes and cellular components related to neurodevelopment and neuronal function. This is consistent with previous reports that have found that loci shared between schizophrenia and intelligence implicated genes involved in neurodevelopment, synaptic integrity, and neurotransmission.^11^

A further aim of the study was to understand the relationship between genetic liability to broad dimensions of cognitive function and schizophrenia symptoms. We calculated PGS for each latent cognitive factor and assessed their relationship with schizophrenia as well as positive symptoms, negative symptoms, and general psychopathology in individuals with schizophrenia. We found that the visuospatial factor PGS and verbal analytic factor PGS significantly predicted schizophrenia but that there were no significant associations between any latent cognitive factor PGS and schizophrenia symptoms. Previous research has consistently demonstrated a significant relationship between PGS for general cognitive ability and schizophrenia however, findings on the relationship between PGS for specific cognitive abilities and schizophrenia are mixed.^49,50^ The lack of consistent results may be due to differences in the methods of assessing and defining cognitive abilities or domains and a consensus on the measurement of cognitive domains would improve comparability across studies. The lack of association between PGS for the latent cognitive factors and schizophrenia symptoms is in keeping with results from a previous study which found no association between an intelligence PGS and positive, negative, and disorganized symptoms in schizophrenia.^14^ These findings suggest that there is distinction in the genetic determinants of cognitive abilities and other symptoms of schizophrenia and extend findings from phenotypic analyses that show minimal association between positive and negative symptoms and cognitive impairment in schizophrenia.^6^

The results of the current study should be interpreted in the context of several limitations. First, the cognitive assessments from the UKB are brief and bespoke. While previous studies have demonstrated that the psychometric properties for most assessments are adequate^51,52^, the findings of the current study require replication in other samples with psychometrically valid measures of cognitive function. Second, the sample sizes and heritability estimates for the cognitive traits differed and differences in power for the univariate GWAS defining each factor may have affected the results of the multivariate GWAS for each factor. Third, the mean scores for the subscales of the PANSS were relatively low, which is expected given that the TOP study limited enrolment to individuals with the capacity to provide informed consent. As the PANSS assesses current symptom severity, a lifetime measure of schizophrenia symptoms may have been more appropriate for assessing the relationship between cognitive abilities and schizophrenia symptoms. Lastly, we restricted our analyses to individuals of European ancestry and our results may not be generalizable to other populations. Efforts to improve the representation of diverse global populations in genomic studies are ongoing^53,54^ and once the relevant large scale datasets for non-European populations become available, the generalizability of the results from the current study should be examined.

In summary, we estimated three genetically determined correlated cognitive factors and applied a variety of genomic methods to explore the relationship between the cognitive factors and schizophrenia. We found extensive polygenic overlap between the latent cognitive factors and schizophrenia and demonstrated that most shared common genetic variants have opposite directions of effect on cognitive abilities and schizophrenia risk. This study demonstrated that most loci shared between the latent cognitive factors and schizophrenia show unique patterns of association with each cognitive factor. Results from biological annotation of shared loci converged and implicated biological processes related to neurodevelopment and neuronal functioning. Lastly, we extended current knowledge with our polygenic risk score analyses which showed a distinction in the common genetic determinants of cognitive abilities and schizophrenia symptoms. Collectively, our results suggest that heterogeneity in the extent of cognitive impairment observed across cognitive domains in schizophrenia reflects differences in genetic risk sharing between specific cognitive domains and schizophrenia.

## Data Availability

All data produced in the present study are available upon reasonable request to the authors

## Acknowledgements

We would like to thank the participants and members of the research teams involved in the UK Biobank and TOP Study. The current study was supported by National Institute of Mental Health (NIMH: Grant number U01MH125053), and The Research Council of Norway (275054). This research has been conducted using data from UK Biobank, a major biomedical database (Project ID number 27412; www.ukbiobank.ac.uk). This work was performed on the TSD (Tjeneste for Sensitive Data) facilities, owned by the University of Oslo, operated and developed by the TSD service group at the University of Oslo, IT-Department (USIT). We gratefully acknowledge support from the Research Council of Norway (324499, 324252, 273291, 223273, 248980, 326813), the South-East Norway Regional Health Authority (2019-108, 2022-073), European Economic Area and Norway grants (no. EEA-RO-NO-2018-0573), KG Jebsen Stiftelsen (SKGJ-MED-021), and the South African Medical Research Council. The content is solely the responsibility of the authors and does not necessarily represent the official views of the funders.

## Author Contributions

OW conceptualized the research question, performed the data analysis, and drafted the manuscript with input from AAS and SD. All authors contributed to data interpretation and editing of the manuscript. All authors approved the final version of the manuscript.

## Ethical Standards

This work was approved by the University of Cape Town Human Research Ethics Committee (reference number - 734/2021). The UKB has ethical approval (REC reference number - 11/NW/0382) and is overseen by an Independent Ethics and Governance council. The TOP Study was approved by the Norwegian Scientific Ethical Committee and the Norwegian Data Protection Agency. Informed consent was obtained from participants in the UKB and TOP Study.

