## Supplementary Note for "Genomic Insights into the Shared and Distinct Genetic Architecture of Cognitive Function and Schizophrenia"

### **Supplementary Information**

#### **UK Biobank Cognitive Tests**

Fluid Intelligence (*n* = 140,797). Fluid intelligence was measured the UKB Fluid IQ Test, which assesses verbal and numerical reasoning. Verbal reasoning is tested by a series of verbal analogy problems and numerical reasoning is tested with number sequence problems. The participants are given a maximum of 2 minutes to answer 13 multiple-choice questions. The score is taken as the number of correct answers. The UKB fluid intelligence test has acceptable test-retest reliability (Pearson *r*_12_ = 0.61)^1^ and internal consistency (Cronbach α = 0.62)^2^.

Mean reaction time and reaction time variability (*n* = 434,321). The UKB reaction time test is designed to measure processing speed and is based on a Go/NoGo test. During the test, participants were asked to push a button when the 2 cards displayed on screen were matching. The test consisted of 12 trials, 9 of which contained matching cards. Mean reaction time was calculated, in milliseconds, for the trials containing matching cards and reaction time variability was taken as the intraindividual standard deviation of response time for correct responses to trials with matching cards. Both mean reaction time and reaction time variability scores were multiplied by negative one so that higher scores were indicative of better performance. The UKB reaction time test has demonstrated good internal consistency (Cronbach α = 0.85)^2^, moderate test-retest reliability (Pearson *r*_12_ = 0.55)^1^, and good concurrent validity with well-validated tests of reaction time.^1^

Numeric Memory (*n* = 44,969). The UKB Numeric Reasoning Test is a backward digit span task designed to assess working memory. Participants are shown a two digit number on the screen, the number disappears, and after a brief pause, the participants are asked to recall the number sequence in reverse order. If the participant makes an error, they are asked to recall a different sequence of the same digit length. The sequence becomes one digit longer every time the participant correctly recalls the digit sequence in reverse order. The test ends when a participant fails to correctly recall two trials of the same digit length or if the participant correctly recalls a 12-digit number. The score is taken as the maximum number of digits correctly remembered in reverse order. The UKB numeric memory test shows reasonably good concurrent validity with validated tests of working memory, and adequate test-retest reliability (Pearson *r*_12_ = 0.50)^1^.

Pairs Matching (*n* = 436,853). The UKB Pairs Matching test measures visual memory. Participants are shown pairs of matching cards randomly arranged in a grid on the screen. Participants are then asked to match pairs from memory when the cards are turned face down. Participants complete two trials; one with three card pairs and one with six card pairs. The overall score was the sum of the number of errors made in each trial before all pairs were identified. Scores were multiplied by negative one so that higher scores were indicative of better performance. The UKB pairs matching demonstrated moderate concurrent validity with other tests of visuospatial memory and modest test-retest reliability (Pearson *r*_12_ = 0.41)^1^.

Prospective Memory (n = 144,031). The UKB Prospective Memory is designed to assess prospective memory, the ability to remember to perform intended actions in future.^3^ The participants were asked to remember instructions for a future task at the beginning of the cognitive test battery. The participants were given a score of 0 or 1 based on whether or not they correctly remembered the instructions and completed the task on the first attempt. The test-retest reliability of UKB Prospective Memory was moderate (Pearson *r*_12_ = 0.45)^1^.

Matrix Pattern Completion (*n* = 28,922). The UKB Matrix Pattern Completion measures non-verbal reasoning and is adapted from the COGNITO Matrices Test^4^. Participants were asked to complete 15 puzzles that ranged in difficulty. For each puzzle, the participant was shown a matrix design with a missing piece, and they were asked to use logic to identify the missing piece from 6-8 options. The test score was the number of correctly solved puzzles in 3 minutes. The UKB matrices showed good concurrent validity with the COGNITO Matrices Test and moderate test-retest reliability (Pearson *r*_12_ = 0.45)^1^.

Paired Associate Learning (*n* = 34,364). The UKB Paired Associate Learning (PAL) test is based on a version of Paired Associate Learning test from Test the Nation ^5^ and is designed to assess verbal declarative memory. During the learning phase of the test, participants were shown 12 word pairs and were instructed to try to remember the word pairs as they would be asked to recall them at a later stage. Next, the participants completed the UKB matrices. Upon completion of the UKB matrices, participants resumed the UKB PAL. Participants were shown one word from a word pair (the target word) and were asked to select the word that completed the pair from a list of four options. Participants completed 10 trials and the score was the correct trials. The UKB PAL has acceptable test-retest reliability (Pearson *r*_12_ = 0.45) ^1^ and good concurrent validity.

Symbol Digit Substitution (*n* = 28,949). UKB Symbol Digit measures processing speed. During the test, participants were shown a key, which paired symbols with numbers. Participants were instructed to use the key to pair a row of symbols with the corresponding number. The score was the number of correct symbol-digit matches made in 60 seconds. The psychometric properties of the UKB Symbol Digit are good.

Tower Rearranging (*n* = 28,688). The UKB Tower Test was adapted from the One-touch Tower of London test ^6^ and is designed to assess planning ability, an aspect of executive function. During each trial of the task, participants were shown a display (display A) of 3 different coloured hoops arranged on 3 pegs. Beneath display A was display B, which also showed 3 different coloured hoops arranged on 3 pegs however, the coloured hoops were arranged in a different order to those shown in display A. For each trial, participants were asked the minimum number of moves required to change display A into display B. The score was the number of correct trials completed in 3 minutes.

Trail Making Test (Part A *n* = 28,899; Part B *n* = 28,156). The UKB Trail Making Test (UKB TMT) is an adapted version of the Halstead-Reitan Trail Making Test^7^, a measure of executive function. During part A of the test, participants were shown the numbers 1-25 pseudo-randomly arranged on the computer screen. Participants were instructed to touch the numbers in ascending order. During part B, the numbers 1-13 and letters A-L were displayed pseudo-randomly on the screen. Participants were instructed to alternate between touching the numbers in ascending order and the letters in alphabetical order. The score was the sum of the time, in deciseconds, taken to complete each part. For this analysis, scores were multiplied by negative one so that higher scores were indicative of better performance.

#### **The Norwegian Thematically Organised Psychosis Research (TOP) Study**

The Norwegian TOP study is an ongoing case-control study that began in 2002. Cases are individuals with a DSM-IV diagnosis of schizophrenia, schizoaffective, and schizophreniform disorder or bipolar disorder I, bipolar disorder II or bipolar disorder not otherwise specified.^8^ Participants are considered eligible if they are aged between 18-65 years and demonstrate the ability to provide written informed consent. Individuals with pronounced cognitive deficits (IQ below 70), severe somatic illness, and brain damage are excluded from the study. Cases are recruited from psychiatric inpatient and outpatient units at major hospitals in the Oslo area, as well as Trondheim, and Southeast regional hospitals in Norway (Sykehuset Innlandet, Sykehuset Østfold). Healthy controls are randomly selected from statistical records of individuals in the same catchment area as cases. Informed consent is provided by all participants and the human subjects protocol was approved by the Norwegian Scientific-Ethical Committee and the Norwegian Data Protection Agency.

The PGS analyses were conducted using a subsample of participants of European ancestry from the TOP study. The subsample of participants included 311 individuals with schizophrenia and 1060 controls. Symptom severity for participants with schizophrenia was assessed using the Positive and Negative Syndrome Scale (PANSS) which has demonstrated good interrater reliability (Intraclass Coefficient = 0.82) in the TOP study.^9^

DNA was extracted from blood and saliva samples collected at enrolment. Genotyping was performed using the Human Omni Express-24 v.1.1 (Illumina Inc., San Diego, CA, USA) at deCODE Genetics (Reykjavik, Iceland). Pre-imputation quality control was performed using PLINK 1.9^10^ and involved removal of SNPs with genotyping rate < 95%, Hardy-Weinberg disequilibrium test p-value < 10-4, high rate of Mendel errors in eventual trios or significant (False Discovery Rate < 0.5) batch effects. Whole individual genotypes were excluded if they had low coverage (< 80%) or high likelihood of contamination (heterozygosity > 5 standard deviations above the mean). The quality-controlled genotypes were phased using Eagle^11^, and missing variants were imputed with MaCH^12,13^ using version 1.1 of the trans-ethnic reference sample put together by the haplotype reference consortium (HRC)^14^. High quality variant sets from the quality control procedure were selected to compute individual’s genetic principal components representing loadings along the 20 first eigenvectors of the pairwise genetic covariance matrix of a sub-sample of unrelated individuals from the HRC panel. Following the quality control and imputation procedure, variants with information score < 0.8 or minor allele frequency lower than 0.01 were removed. In addition, individual genotypes imputed with < 75% confidence were set to missing, the remaining ones were converted to best guess hard allelic dosages.

### **Supplementary Figures**

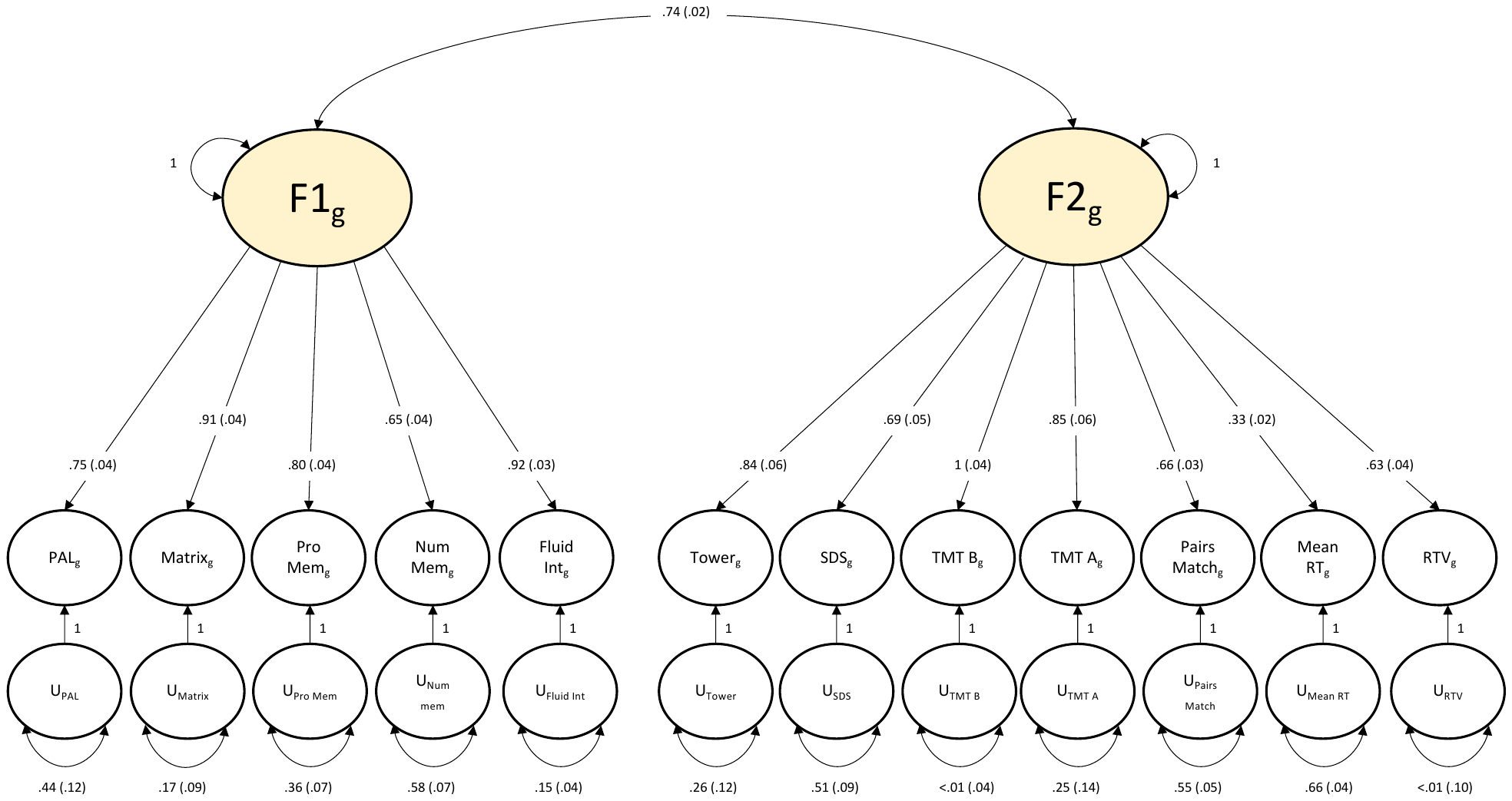

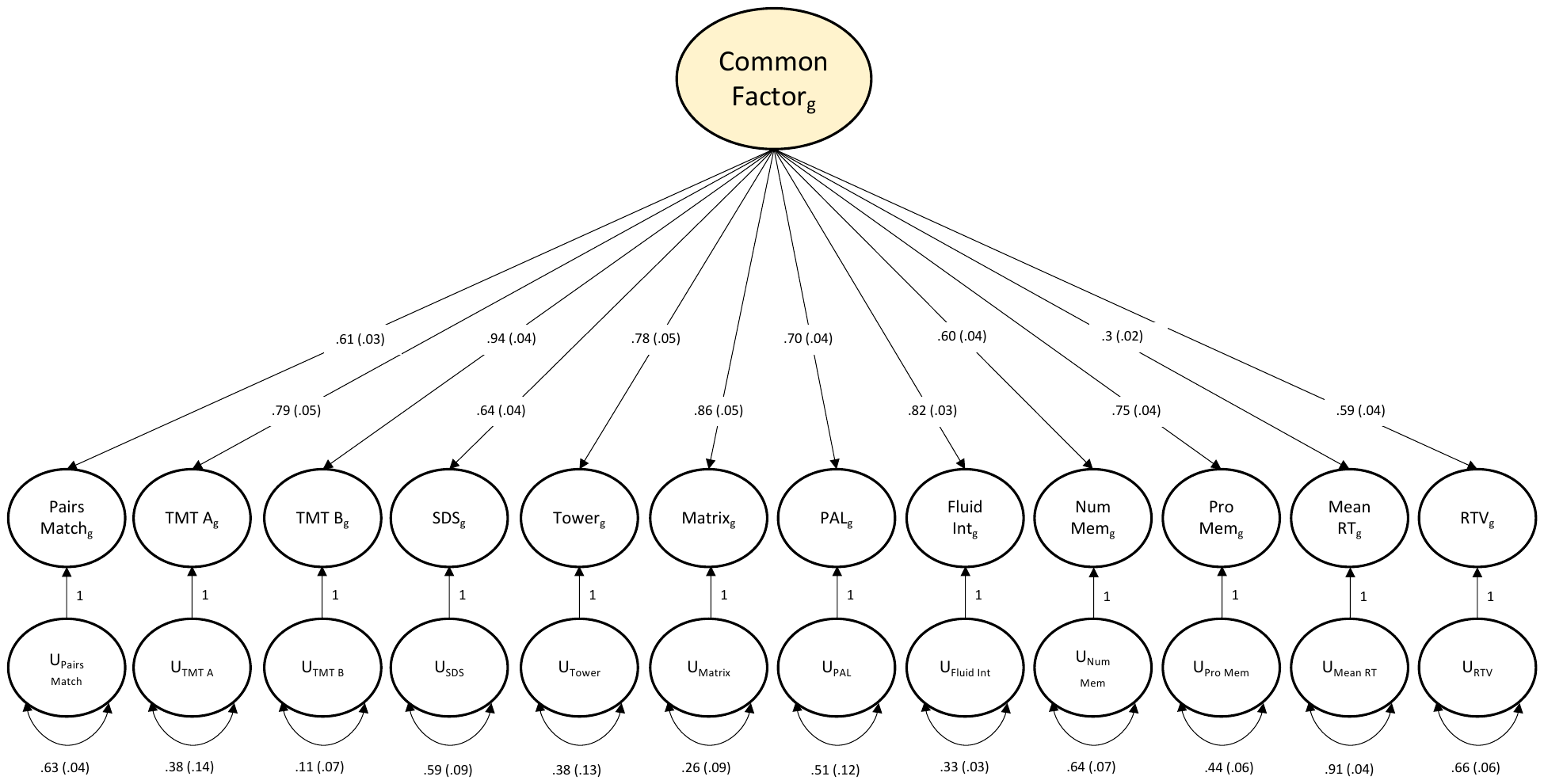

**A**

**B**

**Supplementary Figure 1.** Standardized genetic factor solutions for the covariance structure of twelve UKB cognitive traits. (**A**) The path diagram for the common factor model. (**B**) The path diagram for the correlated two factor model. Circles represent latent variables that are inferred from the data. Single-headed arrows depict regression relationships with the arrows pointing from the independent variables to the dependent variables. Two-headed arrows represent covariance relationships between variables or the residual variance of a variable if the arrow connects the variable to itself. Standard errors of the estimates are in parenthesis.

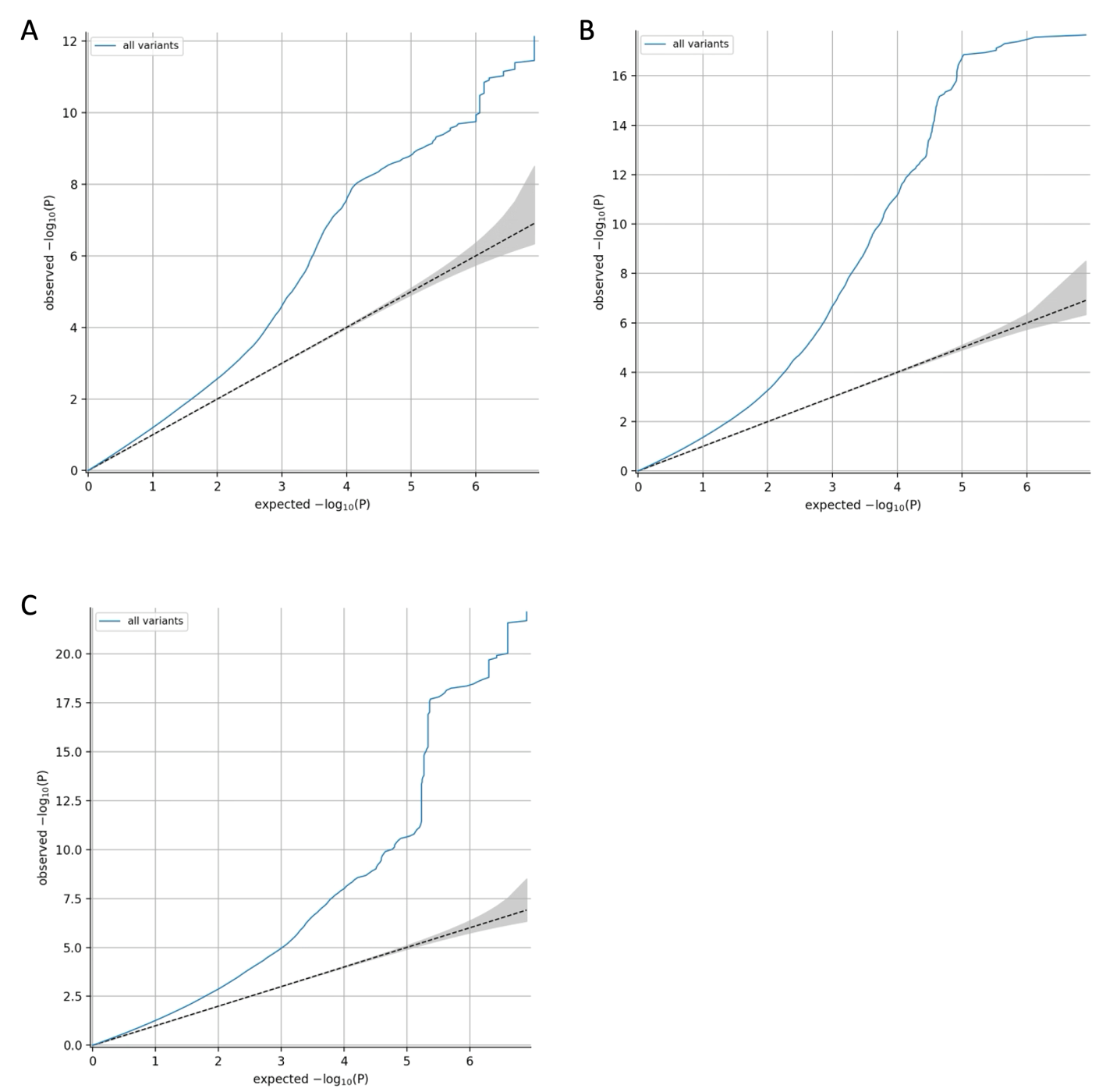

**Supplementary Figure 2.** Quantile-quantile plot of expected under null (no association, x axis) versus observed (y axis) -log10 p-values for the multivariate GWAS of latent cognitive factor 1 (**A**), factor 2 (**B**), and factor 3 (**C**).

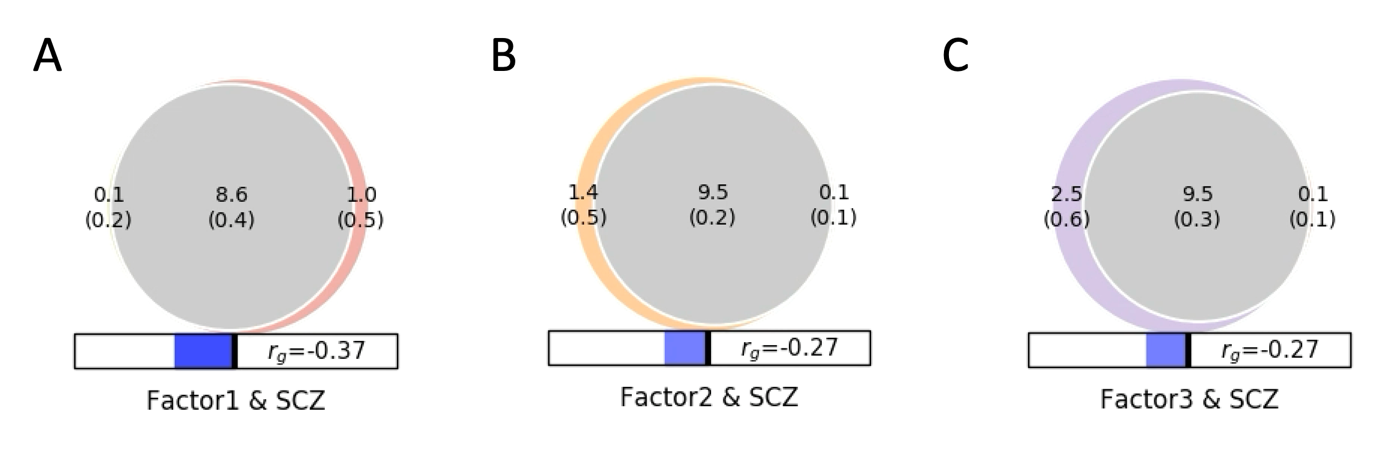
**Supplementary Figure 3.** Bivariate MiXeR results. (**A**) Polygenic overlap between latent cognitive factor 1 (visuospatial processing) and schizophrenia. (**B**) Polygenic overlap between latent cognitive factor 2 (verbal analytic reasoning) and schizophrenia. (**C**) Polygenic overlap between latent cognitive factor 3 (decision/reaction time) and schizophrenia. Venn diagram shows the number (in thousands) of estimated causal variants shared between both traits (grey) and unique to each trait (in colour). Standard deviations of the estimates are shown in parenthesis. The size of the circle represents the extent of polygenicity of each trait.

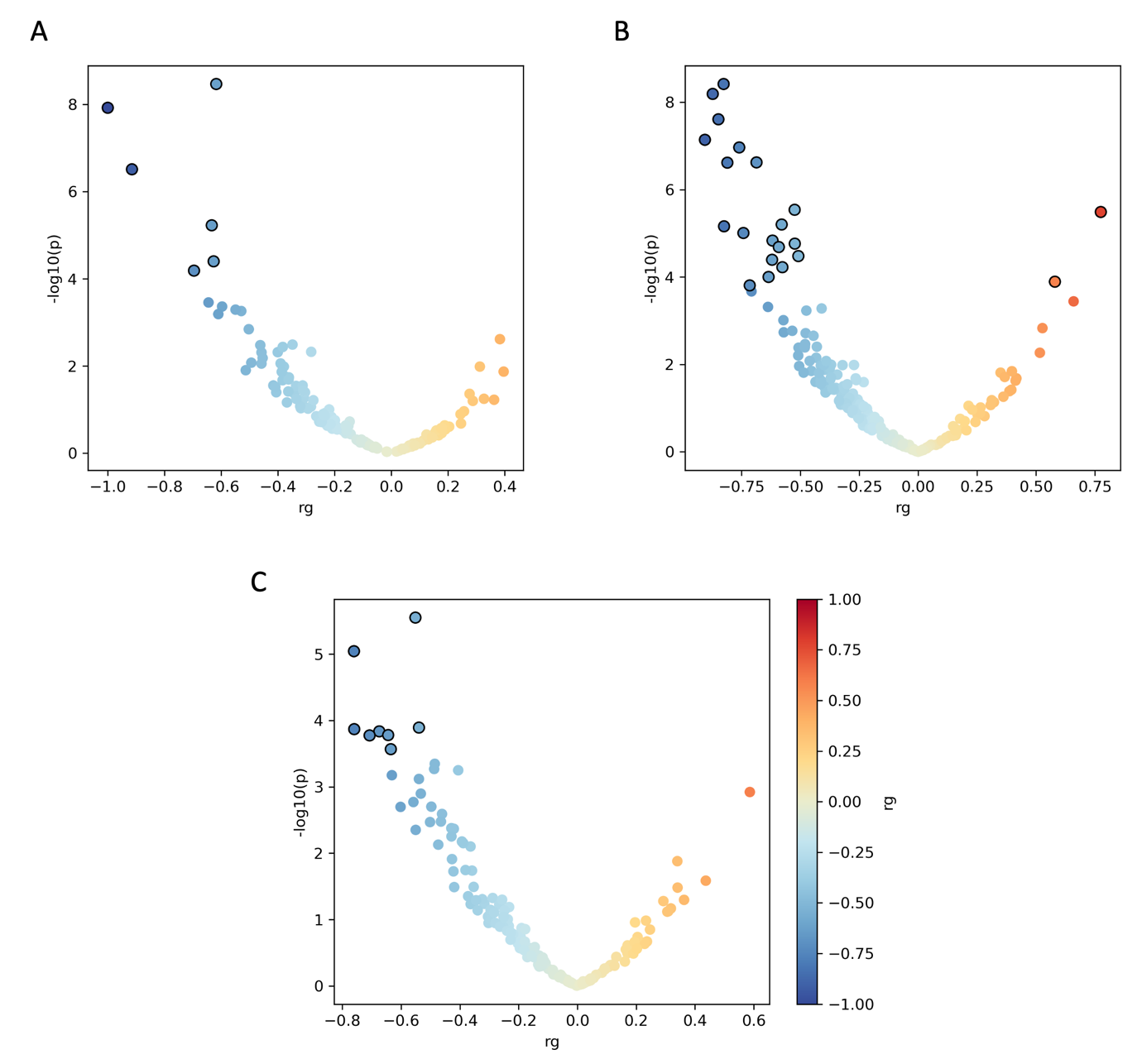

**Supplementary Figure 4.** Volcano plots showing local genetic correlations between schizophrenia and latent cognitive factor 1 (**A**), factor 2 (**B**), and factor 3 (**C**). Local genetic correlations for regions with significant heritability for schizophrenia and the latent cognitive factor are represented as a single point. Regions with a significant genetic correlation after Bonferroni correction are outlined in black.
